# Overcoming Barriers to Warfarin Patient Self-Management (PSM) in the US Healthcare System: Implementation Trial Study Protocol

**DOI:** 10.1101/2025.08.18.25333912

**Authors:** Heeseung Hong, Aaron S. Wilson, Aubrey E. Jones, Sara R. Vazquez, Spencer Gilbert, Daniel C. Malone, Nathorn Chaiyakunapruk, Jordan B. King, Geoffrey D. Barnes, Katelyn W. Sylvester, Gina Dube, Nicole V. Irving, Linh Chan, Bishoy Ragheb, Thomas Delate, Daniel M. Witt

## Abstract

**Introduction:** Warfarin is a narrow therapeutic index drug that requires frequent monitoring using the international normalized ratio (INR). Current clinic-based INR monitoring models lead to suboptimal warfarin management. Warfarin patient self-management (PSM) has consistently demonstrated superior efficacy compared to clinic-based management but is virtually unused in the US healthcare system. The objective of this study is to implement PSM in the US healthcare system using strategies developed to overcome previously identified barriers associated with PSM underutilization as well as potential PSM facilitators.

**Methods and analysis:** We aim to implement PSM with 150 adult patients at four sites using strategies developed to address barriers to PSM specific to the US healthcare system. Implementation strategies will be guided by the Consolidated Framework for Implementation Research and the Quality Implementation Framework supported by Rapid Cycle Research Methodology. A type III hybrid implementation-effectiveness study design will be used to assess PSM implementation strategy outcomes while also gathering information on PSM clinical outcomes centered on the five elements of the RE-AIM framework (Reach, Effectiveness, Adoption, Implementation, Maintenance). This study will be approved by the ethics boards at all participating sites.

**Discussion:** We plan to disseminate the results of this research program examining the feasibility of PSM in US anticoagulation management services in scientific journals and conferences, as well as making elements of the PSM implementation toolkit publicly and freely available.

Trial Registration: ClinicalTrials.gov Identifier: NCT04766216

## Background

Warfarin is a cornerstone of effective and affordable thromboembolism prevention and treatment, accounting for 8 million prescriptions in the United States (US) annually.^1^ Direct oral anticoagulants (DOACs) have emerged as first-line therapies for many indications, such as stroke prevention in atrial fibrillation (AF)^2^ and venous thromboembolism (VTE) treatment.^3^ However, DOACs have been shown to be less effective than warfarin in important patient populations such as prosthetic heart valve replacement,^4^ valvular atrial fibrillation,^5^ and antiphospholipid antibody syndrome.^6^ Additionally, higher costs for DOAC therapy may lead many patients in the US to opt for warfarin therapy. Thus, warfarin therapy will remain a vital oral anticoagulant therapy option for the foreseeable future for many patients.

In the US, clinic-based care is the de facto standard of care for warfarin management. In this care model, patients visit a laboratory or clinic for international normalized ratio (INR) testing and clinicians make warfarin dosing adjustments based on the results.^7^ An alternative model to this burdensome clinic-based approach is patient self-management (PSM) where patients test their INR at home using a portable INR testing device and independently make warfarin-dosing adjustments.^8^ Use of this model has led to clinical outcomes favoring PSM over clinic-based care: a notable 50% reduction in thrombosis and mortality, and improvement in INR control, as well as increased patient satisfaction and quality of life.^8^ Based on this evidence, guidelines from the American College of Chest Physicians,^9^ American Society of Hematology,^10^ and European Society of Cardiology^11^ recommend PSM over other warfarin management strategies. Unfortunately, PSM is virtually nonexistent in the US despite superior outcomes and evidence-based recommendations favoring its use.

Our long-term objective is to overcome barriers to the use of PSM in the US healthcare system. The purpose of this paper is to describe the protocol of a research program focused on increasing the use of PSM in the US. Key elements of the program include developing a warfarin PSM implementation toolkit and evaluating PSM implementation and clinical outcomes through prospective clinical implementation with 6-months of follow-up in four US anticoagulation management services. The conceptual frameworks supporting potential PSM implementation strategies are described initially, then a process for developing a PSM implementation toolkit is outlined, and finally clinical implementation methods are detailed.

### Conceptual Frameworks

PSM implementation strategies for this study will be guided by the Consolidated Framework for Implementation Research (CFIR),^12^ the Quality Implementation Framework (QIF),^13^ and Rapid Cycle Research Methodology (RCRM).^14^

In accordance with QIF Phase 1, which focuses on the implementation setting,^13^ we recruited four anticoagulation management services that had achieved designation as Anticoagulation Centers of Excellence (ACE) by the Anticoagulation Forum to serve as implementation sites for this proposal. Chosen sites include the University of Utah, the University of Michigan, Brigham and Women’s Hospital, and the Veterans Administration Loma Linda Healthcare System.

As specified in QIF Phase 2, a structure for PSM implementation will be developed.^13^ During regular online meetings, investigators and PSM implementation site teams will use a stepwise process to develop an organized structure to oversee the PSM implementation process (Table 1). The organized structure to guide the PSM implementation process will consist of core implementation elements as summarized in Figure 1.

**Table 1.**
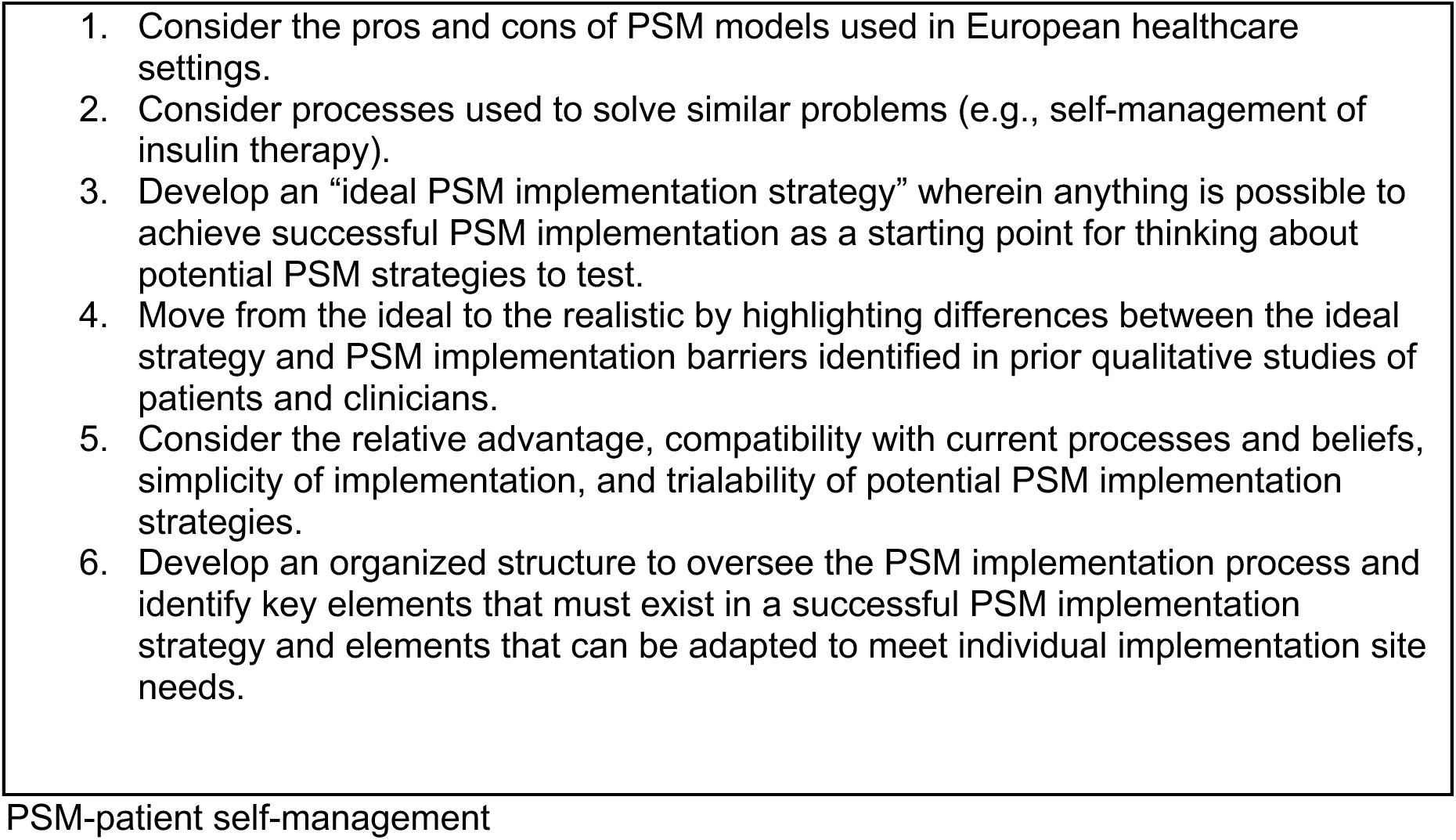
Stepwise process for developing a warfarin PSM implementation strategy

**Figure 1.**
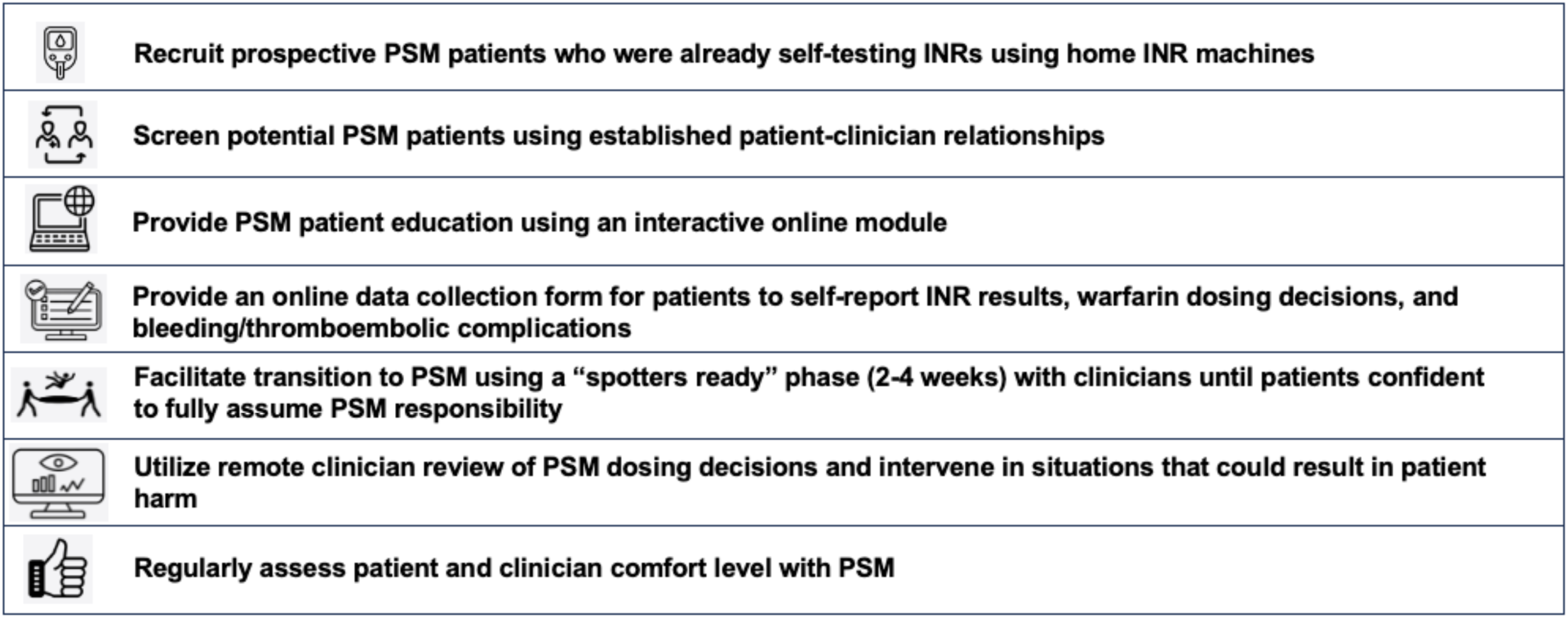
Core elements of the organized structure to guide warfarin patient self management implementation. PSM-patient self management; INR-international normalized ratio.

As specified in QIF Phase 3,^13^ once sites begin transitioning patients to PSM, initial implementation strategies will be refined using the Plan-Do-Study-Act (PDSA) process as described by the Institute for Healthcare Improvement (IHI).^15^

### PSM Implementation Toolkit Development

In accordance with RCRM Phases 3 and 4 (knowledge exploration and solution development),^14^ workgroups will be formed to develop a PSM implementation toolkit with information pertaining to the following areas.

#### Patient selection criteria

General PSM patient selection criteria will include 1) medical stability at the time of PSM initiation; 2) cognitive stability and capable of following instructions and/or has a willing and capable caregiver; and 3) currently self-testing with a home INR device. Specific PSM patient selection criteria related to participation in the PSM implementation study will include 1) goal INR range of 2.0 to 3.0 or 2.5 to 3.5; 2) completed more than 9 months of warfarin therapy at time of enrollment; 3) willing to use single warfarin tablet strength; 4) planned duration of warfarin therapy of at least 6 months; 5) willing to test INR every 7-14 days; 6) receiving majority of care within the enrolling site’s healthcare system; and 7) able to maintain internet access throughout study participation.

#### PSM educational tools and knowledge assessment

PSM educational tools and knowledge assessments will be provided using an interactive online module. Upon conclusion of the online module, patients will complete a PSM knowledge assessment and asked to indicate their PSM comfort level (very uncomfortable, somewhat uncomfortable, somewhat comfortable, very comfortable).

#### Approaches for documenting PSM activities in the EHR

During the PSM implementation phase, participants will self-report INR results, factors that might influence INR results, warfarin dosing decision details, and bleeding or thromboembolic events using an online survey hosted in REDCap (see supplementary material).^16,17^ A summary of the REDCap survey will be emailed to the site study team for review and intervention in situations that could potentially result in harm. PSM INR results will be documented in the participant’s EHR.

#### Participant clinical decision support tools

Participant clinical decision tools for the PSM implementation toolkit will consist of an online dosing tool (“My Warfarin”) and a manual dosing method (“DIY dosing”). Participants or their caregivers will also be allowed to make warfarin dosing decisions based on their own experience without using formal clinical decision support tools if desired. Participants will be instructed to contact their anticoagulation clinician in high-risk situations (e.g., INR <1.5 or >5.0, any bleeding or thromboembolic symptoms, prior to scheduled invasive procedures) or if they want to consult with a clinician for any reason.

### Clinical Implementation Methods

#### Study Design

We will use a type III implementation-effectiveness hybrid research design, which allows evaluation of implementation strategy outcomes while observing and gathering information on PSM’s impact on relevant clinical outcomes.^18^ The five elements of the RE-AIM framework (Reach, Effectiveness, Adoption, Implementation, Maintenance) will be used as an overarching construct for reporting implementation and effectiveness outcomes for this study (Figure 2).^19^ The IRBs of all participating sites gave ethical approval for this work.

**Figure 2.**
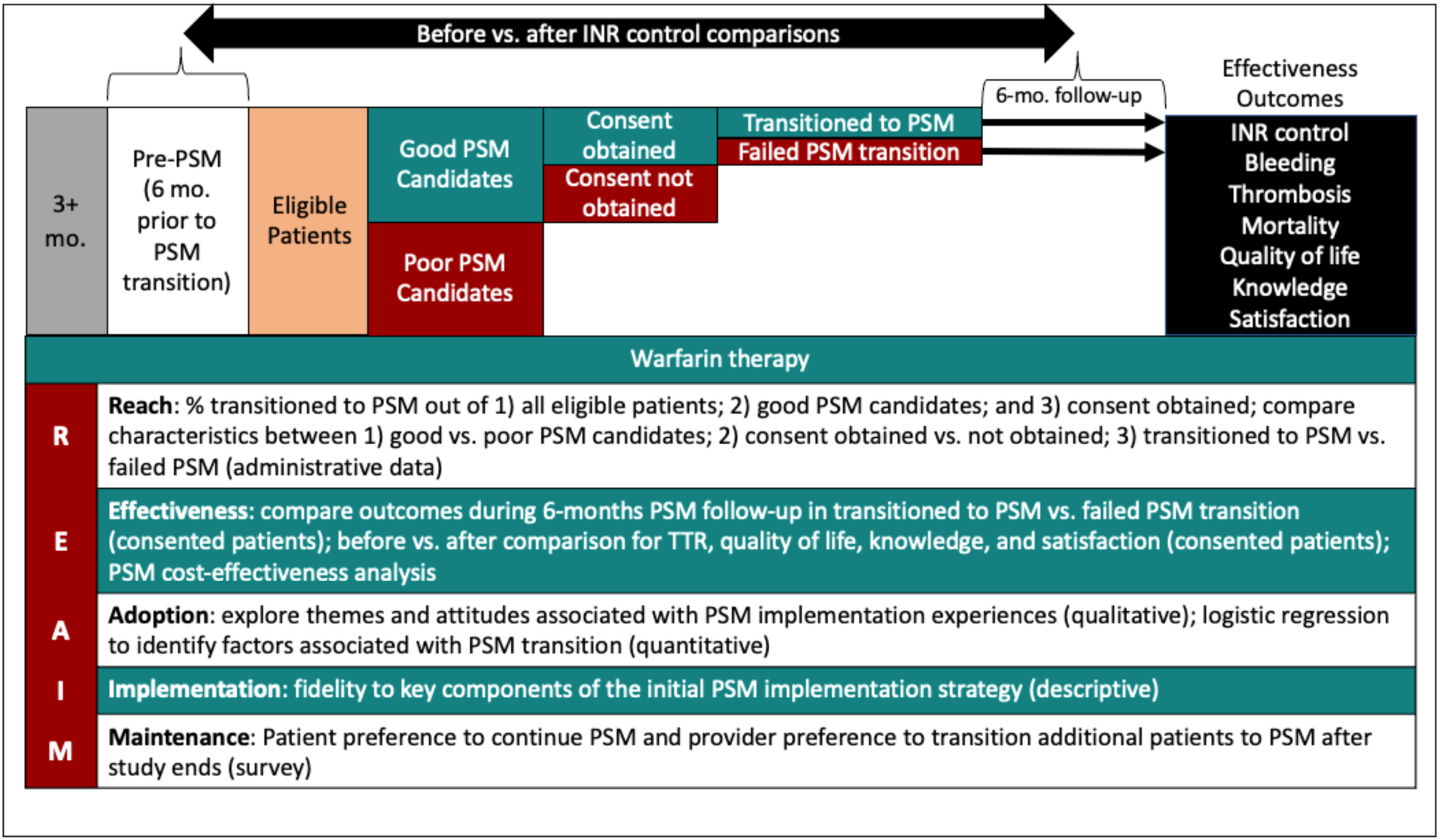
Overview of type III implementation-effectiveness hybrid research design. PSM-patient self-management; INR-international normalized ratio; TTR-time in therapeutic INR range

#### Inclusion and exclusion criteria

Each enrolling site will recruit English-speaking patients at least 18 years of age who meet the previously defined patient selection criteria (see “Patient selection criteria” section above). Each site will consent and enroll eligible patients, track PSM performance, and prospectively collect outcome data using online case-report forms hosted in REDCap.

#### Recruitment process

During usual patient care activities, clinicians will ask patients who meet inclusion criteria if they would be interested in participating in the PSM implementation study. Those expressing interest will be asked a set of screening questions to determine suitability for study participation (Table 2). Clinicians will then record answers to the following questions using REDCap: 1) How confident do you feel in this patient’s ability to perform PSM safely? (Very confident / Somewhat confident / Not confident); 2) Based on the patient’s responses to the screening questions, would you recommend they be included in the PSM implementation study? (Yes/No); 3) Why do you feel this patient is not a good candidate for PSM? (Short answer if applicable). Patients providing satisfactory responses to the screening questions will be forwarded to site study coordinators to discuss further participation in the study and complete the written consent process. Enrollment began March 1, 2023. The timing of study procedures is summarized in Table 3.

**Table 2.** Screener questions for patients potentially interested in warfarin PSM study participation

|  |  |
| --- | --- |
| 1. | Why are you taking warfarin? |
| 2. | What is your INR goal range? |
| 3. | What is your warfarin tablet strength? |
| 4. | What would you do with your warfarin dose if your INR result was too low? |
| 5. | What bad things could potentially happen if your INR gets too high? |

PSM-patient self-management; INR-international normalized ratio

**Table 3.**
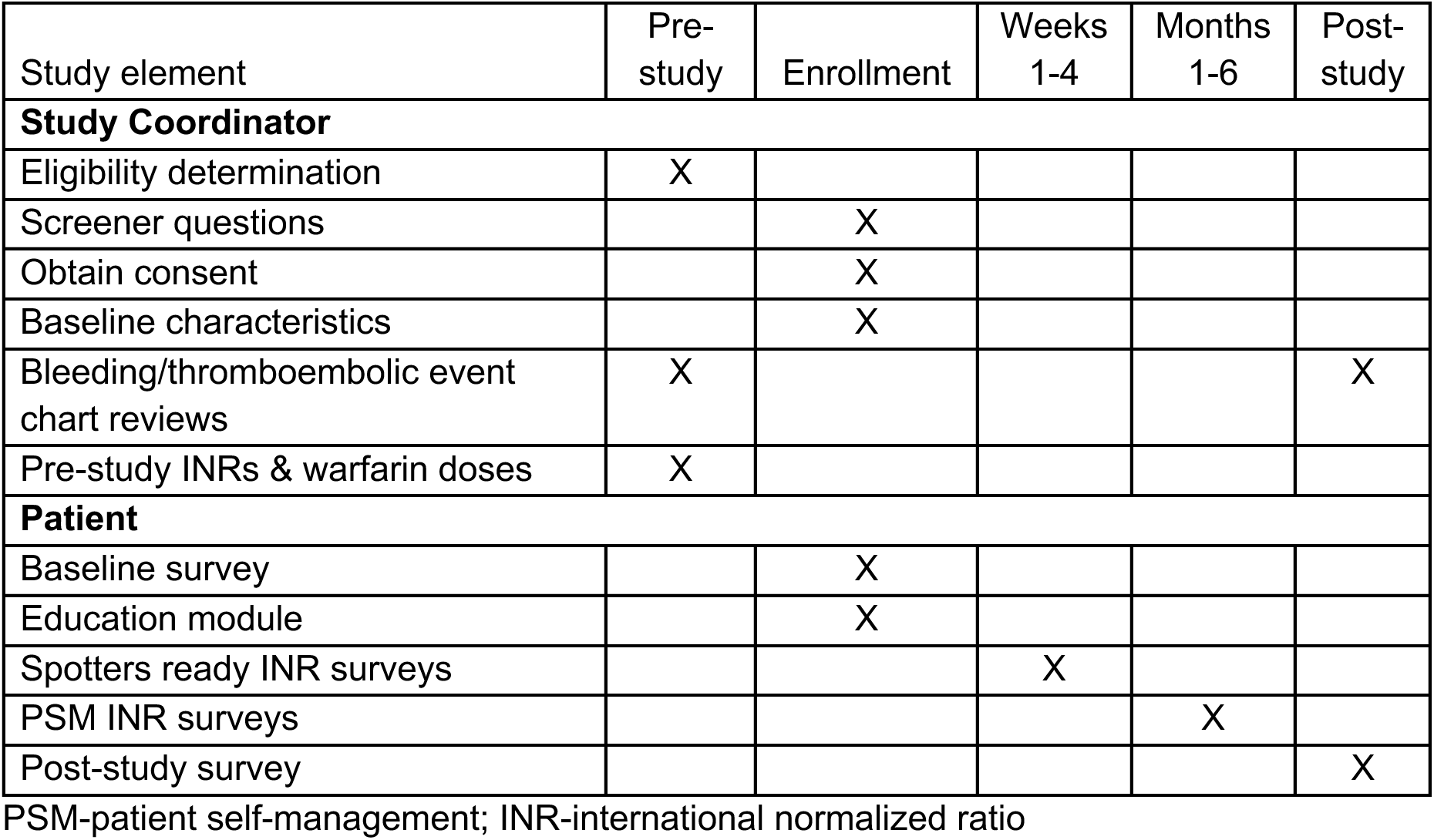
Timing of PSM study data collection

#### PSM implementation phase

Prior to initiating PSM, consenting participants will complete an online survey hosted in REDCap containing the following: 1) the anticoagulation knowledge tool (AKT), a validated 28-item instrument that assesses participants’ anticoagulation knowledge;^20^ 2) the anti-clot treatment scale (ACTS), a 15-item participant-reported instrument assessing satisfaction with anticoagulant treatment that includes a 12-item Burdens scale and a 3-item Benefits scale;^21^ and 3) the Short Form 36 questionnaire (SF-36), a validated instrument used to estimate health-related quality of life.^22^

Participants will complete the previously described interactive online educational module in preparation for initiating PSM. Clinician study team members at each site will use the combination of each participant’s PSM knowledge assessment score, comfort level with PSM, and their subjective assessment of the participant’s likelihood of success with PSM to determine if they should progress to the “spotters ready” phase where clinicians will provide feedback on the participant’s warfarin dosing plans as needed. The “spotters ready” phase will end and the formal PSM phase begin once participants express feeling comfortable starting PSM on their own.

During the 6-month PSM phase, participants will be asked to check INRs every 1 to 2 weeks when INRs are in range and weekly when INR are out of range. Participants will be asked to use clinical decision support tools from the implementation toolkit (My Warfarin, DIY) or use their own experience to make independent warfarin dosing decisions. Participants will be asked to contact clinicians in high-risk situations as previously described or when additional advice is desired.

During the PSM phase, participants will self-report INR results, warfarin dosing decisions, the method used to arrive at dosing decision (i.e., My Warfarin, DIY, or using their own judgement), factors potentially impacting INR response (i.e., changes in other medications, diet, and/or health status), and bleeding/ thromboembolic complications using the previously described online data collection form hosted in REDCap.

At the end of the PSM phase, participants will be asked to complete a final survey consisting of the same questions as the initial survey (AKT, ACT, and SF-36) and additional questions regarding which clinical decision support tools they used (if any) and preference for continuing PSM after study conclusion.

### Outcomes and Analysis

The five elements of the RE-AIM framework will be used as an overarching construct for reporting outcomes for this study (Figure 2).^19^

#### Reach

The primary implementation outcome (the percentage of consented participants successfully transitioned to PSM) will be calculated using the following denominators 1) all patients receiving warfarin; 2) all patients invited to participate in the study and attempt PSM including those who decline participation; and 3) all participants who provide informed consent and attempt transitioning to PSM. Successful transition to PSM will be defined as completing 6-months of PSM. The timing of PSM transition failures during the 6-month follow-up period will be recorded. Reasons patients refuse to participate in PSM implementation activities and reasons patients failed to successfully transition to PSM during implementation efforts will be recorded and categorized.

Data for “reach” outcomes will be summarized using descriptive statistics. Differences in baseline characteristics between eligible patients who do and do not agree to participate in the PSM implementation phase and between those who do and do not successfully transition to PSM will be compared using the chi-squared test of association (categorical variables), Fisher’s exact test (categorical variables that occurred infrequently) or Student’s t-test (continuous variables). Two-sided p-values <0.05 will be considered statistically significant.

#### Effectiveness

The following INR control outcomes will be compared between eligible participants who do and do not successfully transition to PSM during the 6-month follow-up period, 1) individual time in the therapeutic INR range (TTR) using linear interpolation;^23^ and 2) proportion of INRs resulting in a warfarin dose change. INR control outcomes will also be compared between the 6-month periods before and after PSM transition attempts (Figure 2).

Additional effectiveness outcome comparisons before and after PSM transition will include major bleeding and clinically relevant non-major bleeding events as defined by the International Society on Thrombosis and Haemostasis,^24,25^ thromboembolic events, and all-cause mortality. Thromboembolic events will include stroke, systemic embolism or venous thromboembolism documented by objective imaging. Bleeding and thromboembolic events will be confirmed by manual EHR review. For participants who successfully transition to PSM, AKT, ACT, and SF-36 scores will be compared before and after PSM transition.

Data analysis for “effectiveness” outcomes will consist of basic descriptive statistics. T-tests and the chi-squared test of association will be used to compare differences in continuous and categorical outcomes between patients who do and do not successfully transition to PSM during the 6-month PSM phase. Paired t-tests and McNemar’s test will be used to compare differences between continuous and categorical outcomes before and after PSM transition, respectively. Two-sided P-values <0.05 will be considered statistically significant.

#### Adoption

Both qualitative and quantitative analyses will be used to assess adoption. *Qualitative methodology* will be used to explore themes and attitudes associated with *actual* PSM implementation experiences. A sample of participants who implemented PSM will be invited to participate in individual interviews using a semi-structured interview template. Qualitative analysis of patient interview transcripts will be performed to identify emerging “adoption” outcome themes using an inductive approach. Two investigators will independently review anonymized transcripts line-by-line, annotating passages of text with code to help catalog key concepts. Disagreements between reviewers will be resolved through discussion or by a third reviewer when necessary. Additional participants will be interviewed until it is evident that new themes are unlikely to emerge from further interviews (i.e., saturation). Participant demographic data will be summarized using descriptive statistics. ATLAS.ti, a qualitative data management program (ATLAS.ti Scientific Software Development GmbH, Berlin, Germany), will be used to facilitate the organization, management, coding, and thematic analysis of qualitative data.

*Quantitative methodology* will be used to identify characteristics associated with willingness to participate in the PSM implementation study. Participant characteristics will be entered into multivariable logistic regression models to identify variables that are independently associated with willingness to participate in the PSM phase of the study. The comparator group for this outcome will be patients who declined study participation. Variables collected for each patient will include age at time of the study participation invitation, gender, race, ethnicity, indication(s) for warfarin therapy, duration of warfarin therapy at time of study participation invitation, and warfarin tablet strength being used at time of study participation invitation.

#### Implementation (Fidelity)

Self-reported INR values measured during the 6-month follow up period will be individually assessed for fidelity to key PSM elements as described below. For enrolled participants actively engaged in PSM, the percentage of INRs independently and appropriately managed by the participant (i.e., participant used INR to determine warfarin dosing plan while neither contacting a clinician nor taking actions deemed inappropriate by clinicians) will be calculated. Adherence to key PSM elements will be categorized as adherent or nonadherent for each INR. Data for this outcome will be censured on the date of PSM failure or the study end-date, whichever comes first. INRs tested within ±3 days of the date specified by PSM participants will be categorized as fully adherent and INRs measured >3 days before or after as nonadherent. Adherence to warfarin dose selection recommendations as specified by the My Warfarin online decision support tool will be evaluated for each INR. Warfarin doses within ±5% of the specified dose recommendation based on the INR result will be considered adherent.

For “implementation” outcomes, the percentage of INRs falling into adherent and nonadherent categories will be calculated for each patient using the total number of INRs performed during the PSM phase as the denominator. The mean (standard deviation) percentages for each adherence category will be summarized, stratified by implementation site, and compared using ANOVA. An omnibus P-value <0.05 will be considered statistically significant and multiple comparison tests used to determine if significant differences exist between specific sites if needed.

#### Maintenance

Participants engaged in PSM at the end of follow up will be asked to respond to the following prompt, “I would like to continue PSM following study completion” (Yes, No, or Unsure) and will be given the opportunity to provide comments and/or reasons for their answer.

Clinicians at each site will be asked to state whether the participant should be allowed to continue PSM following study completion (Yes, No, Unsure). For “maintenance” outcomes, concordance for continued PSM preference following study completion between patients and clinicians will be assessed using the kappa statistic. A p-value <0.05 will be considered statistically significant.

## Discussion

This study is designed to answer an important question regarding the feasibility of warfarin PSM implementation in the US healthcare system. While PSM is recommended as the preferred warfarin management strategy by evidence-based guidelines it is virtually unused in the US healthcare system despite being widely used elsewhere in the world.^9–11^ Our study team members previously identified addressable barriers to and potential facilitators of wider PSM implementation from the perspective of US patients receiving warfarin and clinicians who manage them.^26,27^ The results of these earlier qualitative studies were used in the rationale and design of the protocol for this implementation study. Our study is intended to demonstrate the feasibility of PSM implementation in the US healthcare system and generate an implementation toolkit for use by others interested in exploring PSM use at their clinical sites.

This study will be among the largest to assess the feasibility of PSM implementation in US settings and the first to use an implementation toolkit specifically designed to address barriers and incorporate facilitators previously identified through qualitative focus groups and interviews with US clinicians and patients, respectively.^26,27^ To our knowledge only three small studies evaluating PSM in the US have been conducted to date.^28–30^ The planned enrollment of 150 patients for our study will exceed that of all previous US-based studies combined.

The proposed study will provide crucial information on the characteristics of patients likely to be interested in attempting PSM, the performance of PSM patient educational tools, the usability of clinical decision support tools, the capability of US patients to safely engage in PSM, and the acceptability of PSM workflows in US healthcare settings.

We chose the RE-AIM framework as it has been shown to highlight essential program elements including external validity that can improve the sustainable adoption and implementation of effective, generalizable, evidence-based interventions like PSM.^30,31^ Further, we chose to utilize anticoagulation management services that have achieved Anticoagulation Centers of Excellence status as defined by the Anticoagulation Forum based on the premise that successful PSM implementation at these sites will promote dissemination of PSM implementation strategies to other centers throughout the US.

This study is registered on ClinicalTrials.gov (NCT04766216) and will be updated with any modifications and results. The outcomes of the study will be presented at scientific meetings and reported in peer-reviewed journals. The data that support the findings of this study will be available from the corresponding author upon reasonable request. Access will be granted to qualified researchers for the purposes of academic, non-commercial research, in accordance with institutional and ethical guidelines.

## Author’s contributions

**Conceptualization:** H.H., A.S.W., A.E.J., S.R.V., D.C.M., N.C., J.B.K., G.D.B., K.W.S., L.C., T.D., D.M.W.; **Data Curation:** H.H., A.S.W., A.E.J., S.G., D.M.W.; **Formal Analysis:** H.H., S.G., T.D., D.M.W.; **Funding Acquisition:** A.E.J., S.R.V., D.C.M., N.C., J.B.K., G.D.B., K.W.S., L.C., B.R., D.M.W.; **Investigation:** H.H., A.S.W., A.E.J., S.R.V., S.G., G.D.B., K.W.S., G.D., N.V.I., L.C., D.M.W.; **Methodology:** A.E.J., S.R.V., D.C.M., N.C., J.B.K., G.D.B., K.W.S., L.C., B.R., T.D., D.M.W.; **Project Administration:** H.H., A.E.J., S.R.V., G.D.B., K.W.S., L.C., D.M.W.; **Software:** T.D.; **Supervision:** G.D.B., K.W.S., L.C., D.M.W.; **Validation:** H.H., A.E.J., T.D., D.M.W.; **Writing—Original Draft Preparation:** H.H., D.M.W.; **Writing—Review & Editing:** H.H., A.S.W., A.E.J., S.R.V., S.G., D.C.M., N.C., J.B.K., G.D.B., K.W.S., G.D., N.V.I., L.C., B.R., T.D., D.M.W.

## Data Availability

All data produced in the present study are available upon reasonable request to the authors

## Supplementary material

| REDCap survey to be completed by patient with each INR measured during the PSM phase |  |  |
| --- | --- | --- |
| Survey element | Response option(s) | Comment |
| 1. What health system do you use for your warfarin? | -Brigham and Women's Hospital<br>-Loma Linda VAMC<br>-University of Michigan<br>-University of Utah |  |
| 2. What is your study ID? | -Patient enters assigned study ID number |  |
| 3. What is your goal INR range? | -2.0-3.0<br>-2.5-3.5 |  |
| 4. Date of INR being reported? | -Patient enters date INR performed |  |
| 5. Value of INR being reported? | -Patient enters INR value | If INR is 5.0 or higher, patient instructed to contact their anticoagulation provider |
| 6. Based on your INR value, do you plan on adjusting your weekly warfarin dose? | -Yes<br>-No | If "No" survey skips to question 20 |
| 7. In response to this INR, who ultimately decided the new warfarin dose you will take? | -I did<br>-Someone else/healthcare provider<br>-Other | If "I did", question 8 is asked; otherwise, survey skips to question 9 |
| 8. How did you decide which warfarin dose to take? | -My warfarin website tool<br>-My own decision<br>-Other |  |
| 9. Over the past week what tablet strength of warfarin did you use? | -1 mg (pink)<br>-2 mg (lavender)<br>-2.5 mg (green)<br>-3 mg (tan)<br>-4 mg (blue)<br>-5 mg (peach)<br>-6 mg (teal)<br>-7.5 mg (yellow)<br>-10 mg (white)<br>-I used different tablet strengths (more than one color) |  |
| 10. Over the PAST week, how many tablets did you actually take each day (enter 0 for missed doses)? | -Patient fills in a grid with the number of tablets taken each day | Dates for the grid will auto populate based on the INR date entered for question 4<br>Total milligrams taken over the previous week automatically calculate from values entered in the grid |
| 11. Since your last INR, which of the following changes have occurred? | -Bleeding symptoms<br>-Clotting symptoms<br>-New or worsening illness | If "Bleeding symptoms" question 12 is asked |
|  | -Changes in medications<br>-Changes in diet/vitamin K intake<br>-None | If “Clotting symptoms” question 13 is asked<br>If “New or worsening illness” question 14 is asked<br>If “Changes in medications” question 15 is asked<br>If “Changes in diet/vitamin K intake” question 16 is asked<br>If “None” survey skips to question 18 |
| 12. What bleeding symptoms (select all that apply)? | -Nosebleed<br>-Unusual bruising (i.e., more severe or more frequent than usual)<br>-Heavy menstrual bleeding<br>-Blood in urine (red or pink)<br>-Blood in stool (bright red or black)<br>-Coughing up blood<br>-Throwing up blood<br>-Stroke symptoms such as facial drooping, arm weakness, and/or speech difficulties<br>-Other |  |
| 13. What clotting symptoms (select all that apply)? | -New pain, redness, or swelling of legs or arms<br>-Chest pain (i.e., more severe or frequent than usual)<br>-Shortness of breath (i.e., more severe or frequent than usual)<br>-Stroke symptoms such as facial drooping, arm weakness, and/or speech difficulties<br>-Other |  |
| 14. What new or worsening illness symptoms (select all that apply) | -Fever or infection<br>-Vomiting (i.e., more severe or frequent than usual)<br>-Diarrhea (i.e., more severe or frequent than usual)<br>-Visit to urgent care<br>-Visit to emergency room<br>-Admitted to hospital<br>-Other |  |
| 15. What changes in medications (prescription, over the counter, and/or | -Started new medication(s)<br>-Stopped medication(s)<br>-Changed dose of medication(s) | Patients asked to provide details of any changes |
| supplements) (select all that apply)? | -Other |  |
| 16. What changes in diet/vitamin K intake (e.g., green leafy vegetables, vegetable smoothies, nutrition, or vitamin supplements with vitamin K) | -INCREASED vitamin K intake<br>-DECREASED vitamin K intake<br>-Other |  |
| 17. Did you contact your anticoagulation management service provider regarding these symptoms? | -Yes<br>-No |  |
| 18. Did you already take your warfarin dose for today before checking your INR? | -Yes<br>-No | Used to auto-populate dates of the grid for question 19 |
| 19. Over the NEXT week, how many tablets do you plan on taking each day? | -Patient fills in a grid with the planned number of tablets for each day | Total milligrams to be taken over the next week automatically calculated from values entered in the grid |
| 20. When do you plan on rechecking your next INR? | -Patient enters date of next INR |  |
| 21. Is there anything else you wanted to tell us about today's INR? | -Text box for patient comments |  |

INR-international normalized ratio; PSM-patient self-management; VAMC-Veterans Administration Medical Center; ID-identification

